# Effects of Various Policy Options on COVID-19 Cases in Nova Scotia including Vaccination Rollout Schedule: A Modelling Study

**DOI:** 10.1101/2021.07.28.21261219

**Authors:** Melissa Gillis, Ahmed Saif, Matthew Murphy, Noreen Kamal

**Author notes:** **Corresponding Author:** Noreen Kamal, 5269 Morris Street, Room 100, PO Box 15000, Halifax NS B3H 4R2. **Funding:** Funding for this project was provided by NSERC Alliance Grant (PI: A. Saif). **Declaration of Authors:** M. Murphy is co-leading the Nova Scotia COVID-19 – Joint Analytics Working Group. All other authors (MG, NK, AS) have no competing interests.

## Abstract

**Background:** The COVID-19 pandemic presents a significant challenge to minimize mortality and hospitalizations due to this disease. Vaccinations have begun to roll-out; however, restriction policies required during and after the rollout remain uncertain. A susceptible-exposed-infected-recovered (SEIR) model was developed for Nova Scotia, and it accounted for the province’s policy interventions, demographics, and vaccine rollout schedule.

**Methods:** A modified SEIR model was developed to simulate the spread and outcomes from COVID-19 in Nova Scotia under different policy options. The model incorporated the age distribution and co-morbidity of the province. A system dynamics model was developed in Vensim. Several scenarios were run to determine the effects of various policy options and loosening of restrictions during and after the vaccine roll-out period.

**Results:** When restrictions policy include moderate closure of businesses, restricting travel to Atlantic Canada, and the mandating of masks and physical distancing, the number of cumulative infections after 110 days was less than 120. However, if national travel was opened by July 5 2021 and there were no restrictions by September 2021, the number of active infections will peak at 6,114 by February 16 2022, and there will be a peak of 104 hospitalizations on February 16 2022. Immediate opening of travel and all restrictions on March 15, 2021 will result in 71,731 active infections by June 4 2021.

**Discussion:** Moderate restrictions will be required even after the population is fully vaccinated in order to avoid a large number of infections and hospitalizations because herd immunity is not reached due to children under 12 not being vaccinated, the efficacy of the vaccine, and the portion of the population that will choose not to be vaccinated.

## INTRODUCTION

Countries around the world are struggling to limit the spread of Corona Virus Disease 2019 (COVID19) since the later part of 2019 and start of 2020. Responding to the effects of COVID 19 is challenging due to the virus’s high infectivity, long incubation period, and acute health impact on the population. The problem is aggravated by the inadequate preparedness of healthcare systems, as current Canadian health systems have been optimized to run at capacity or near capacity. It has been estimated that, in the absence of a proper response, up to 425 million people could be infected and millions could lose their lives because of this pandemic [1].

Several countries have taken extreme actions to contain the spread of the COVID-19 disease. These actions include: travel restrictions, home quarantine, centralized quarantine, and contact tracing [2]. In Nova Scotia, the Provincial Government has declared a State of Emergency that restricts travel, limits gatherings, imposes mandatory self-quarantine for people that have travelled or are exhibiting symptoms, and implemented physical distancing protocols which require a minimum distance of 2 meters between individuals [3]. Similar actions have been taken by numerous other countries to varying degrees of success [4,5]. Nevertheless, all public interventions have societal and economic costs, which include economic losses resulting from economic shut-downs, and the impending negative social impacts of isolation strategies [6]. There are concerns about whether strict policies can be sustained until the population can be vaccinated [7].

Mathematical models are being developed to predict the spread of the virus and its impact on the health care systems and the economy. The Susceptible-Infected-Recovered (SIR) and Susceptible-Exposed-Infected-Recovered (SEIR) models are commonly being used to predict the spread of the COVID-19 and the impact on health care resources [8-12]. Modelling is also being used to determine the impact of policy decision on the spread of the virus [13-15]. We developed a modified, age stratified SEIR simulation model for the province of Nova Scotia that took into account the province’s policy interventions, demographics, and vaccine rollout schedule. Several different policy options were tested to determine their impact on new infections and hospital resources.

## METHODS

We developed a modified SEIR model to simulate the spread and outcomes from COVID-19 in Nova Scotia under different policy options. In the conceptual model (Figure 1), the overall population is considered susceptible to the disease and imported cases are pulled from the susceptible group to maintain a constant population. This is then split into two categories: *exposed* refers to those people who have been exposed and are still moving around the community; *exposed (quarantined)* refers to those people who are exposed but have been identified through contact tracing and are self-isolating according to policy restrictions. Both of these streams move into the infectious period prior to developing symptoms (pre-symptomatic), and then the infectious groups are further split into the severity of the disease. A portion of the severe patients are hospitalized where they can recover or move to the Intensive Care Unit (ICU). Those patients in the ICU can then recover or die. Vaccinations have also been taken into account, and the model considers the efficacy of the vaccine, as well as the proportion of the population that will voluntarily receive a vaccine. The vaccination stream pulls from the susceptible group and moves into the recovered group.

**Figure 1:**
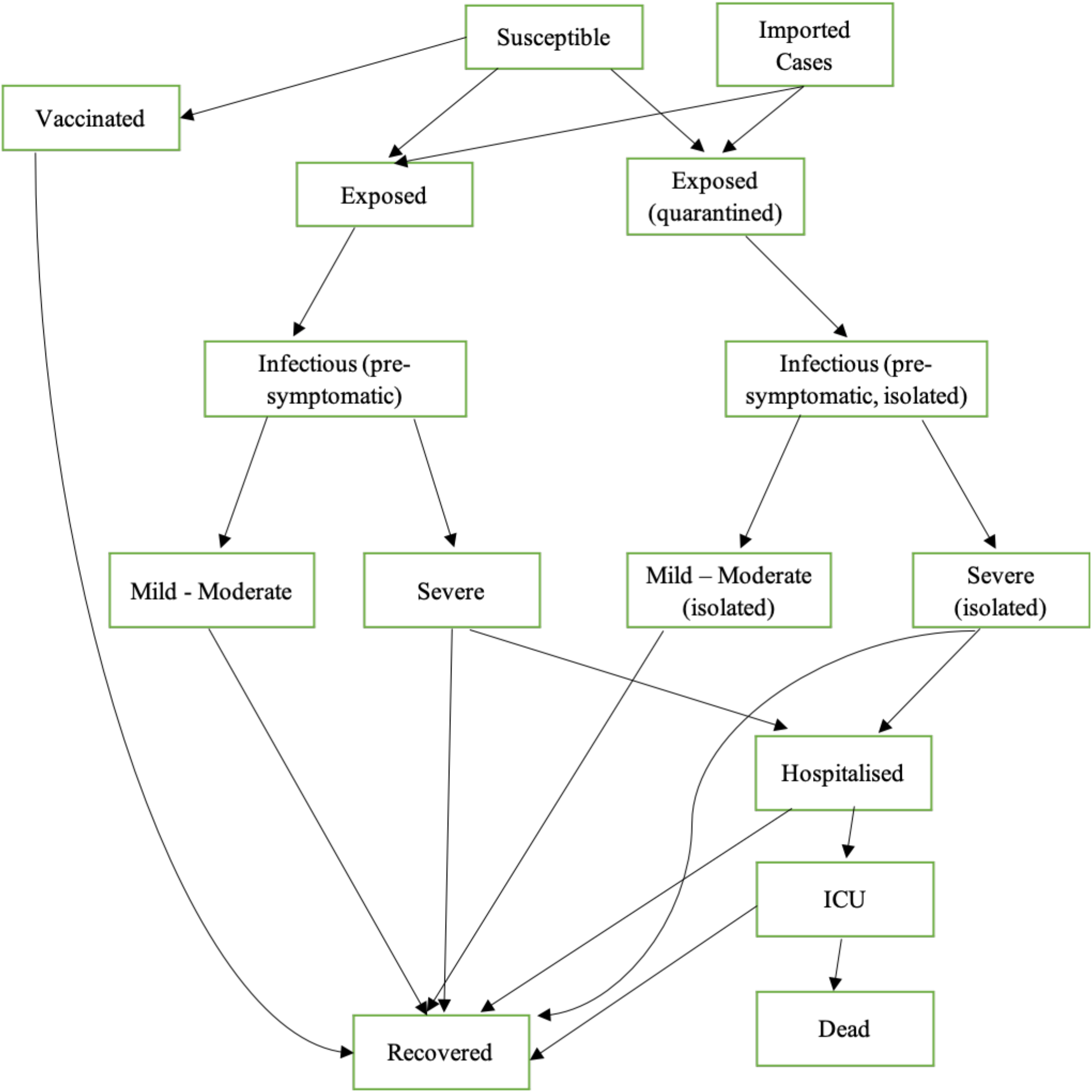
SEIR Model Schematic for Simulating COVID-19 in Nova Scotia. ICU: Intensive Care Unit

### Age Distribution and Co-Morbidities

Nova Scotia’s population is unique in comparison to other Canadian provinces, as the population is older with more co-morbidities [16-18]. This has a significant impact in modelling the COVID-19 pandemic because age and co-morbidities are associated with a greater probability of hospitalization and mortality [19]. The model incorporated the age distribution of Nova Scotia and we dichotomized each group: those with co-morbidities and those without co-morbidities. Table 1 shows the final probabilities that we used in our model for each age group with and without co-morbidities. The probability represents the movement from each level of the SEIR Model shown in Figure 1; for example, in the over 80 years old group without co-morbidities, there is a 35% probability that the infection will be severe, and then from those that are severe, there will be a 26.2% probability of hospitalization, and so forth.

**Table 1:** Probabilities Used for Each Age Group with and without Co-morbidities. CM: Co-morbidities

| *Age Groups* | *Probability Severe* |  | *Probability Hospital* |  | *Probability ICU* |  | *Probability Death* |  |
| --- | --- | --- | --- | --- | --- | --- | --- | --- |
|  | No CM | With CM | No CM | With CM | No CM | With CM | No CM | With CM |
| *0 to 4* | 0.01 | 0.02 | 0.011 | 0.024 | 0.26 | 0.26 | 0 | 0 |
| *5 to 9* | 0.01 | 0.02 | 0.011 | 0.024 | 0.26 | 0.26 | 0 | 0 |
| *10 to 14* | 0.01 | 0.02 | 0.011 | 0.024 | 0.26 | 0.26 | 0 | 0 |
| *15 to 19* | 0.03 | 0.06 | 0.011 | 0.024 | 0.26 | 0.26 | 0.2 | 0.53 |
| *20 to 24* | 0.03 | 0.06 | 0.021 | 0.044 | 0.26 | 0.26 | 0.2 | 0.53 |
| *25 to 29* | 0.03 | 0.06 | 0.021 | 0.044 | 0.26 | 0.26 | 0.2 | 0.53 |
| *30 to 34* | 0.03 | 0.06 | 0.025 | 0.054 | 0.26 | 0.26 | 0.2 | 0.53 |
| *35 to 39* | 0.03 | 0.06 | 0.025 | 0.054 | 0.26 | 0.26 | 0.2 | 0.53 |
| *40 to 44* | 0.03 | 0.06 | 0.035 | 0.075 | 0.26 | 0.26 | 0.2 | 0.53 |
| *45 to 49* | 0.03 | 0.06 | 0.035 | 0.075 | 0.26 | 0.26 | 0.2 | 0.53 |
| *50 to 54* | 0.12 | 0.25 | 0.077 | 0.165 | 0.26 | 0.26 | 0.36 | 0.9 |
| *55 to 59* | 0.12 | 0.25 | 0.077 | 0.165 | 0.26 | 0.26 | 0.36 | 0.9 |
| *60 to 64* | 0.12 | 0.25 | 0.159 | 0.34 | 0.26 | 0.26 | 0.36 | 0.9 |
| *65 to 69* | 0.12 | 0.25 | 0.159 | 0.34 | 0.26 | 0.26 | 0.36 | 0.9 |
| *70 to 74* | 0.35 | 0.76 | 0.262 | 0.561 | 0.26 | 0.26 | 0.58 | 1 |
| *75 to 79* | 0.35 | 0.76 | 0.262 | 0.561 | 0.26 | 0.26 | 0.58 | 1 |
| *80 +* | 0.35 | 0.76 | 0.262 | 0.561 | 0.26 | 0.26 | 0.58 | 1 |

Transmission of COVID-19 is driven by individuals’ social contacts. The typical SEIR framework assumes homogeneous mixing, which does not accurately represent population mixing dynamics. To mitigate the impact, mixing was stratified by age in the model. Mossong et al. [20] determined contact patterns for different age groups to help understand the spread of respiratory diseases. The age to age contact matrix was calibrated using the transmissibility parameter to get an acceptable basic reproduction number (R0) of 3.47 [21], where R0 represents the number of secondary infections cause by a single infected member into a completely susceptible population. Using the calibrated contact matrix, we extrapolated the changes in contact rate for various restrictions that were imposed by the Nova Scotia government.

The full model formulation is provided in the On-Line Supplemental document.

### Policy Restrictions

The policies used in our model were derived by reviewing the policies implemented within Nova Scotia from March through July 2020 [22]. This allowed for review of a large number of policies with varying degrees of strictness. Generally, policies implemented during March and April were stricter, whereas the policies from May through July gradually relaxed restrictions. Examples of restrictions include: closing childcare facilities and, personal services, border closures, shifting to virtual education, and cessation of elective surgeries. The list was consolidated into three overarching categories: closure, protection, and travel. Each category was narrowed down into policies that were assumed to impact infection rate. The main closure components are gathering limitations and closure of schools, universities or colleges, daycare facilities, service businesses, and public outdoor facilities. These policies focus on reducing the average number of daily contacts of an individual. There are five different closure levels with varying degrees of strictness, ranging from keeping everything open to sheltering in place. Protection policies focus on reducing transmissibility, and account for the implementation of physical distancing and masks. Two protection policy alternatives are considered: enforced and not enforced. The travel restriction policies account for the limitations on travellers to prevent case importation. There are four travel policies, where level one is allowing all travel including international and domestic and level four is disallowing all travel. Table 2 shows the policy restrictions used in our simulation model. When the infections began in Nova Scotia, the province had no restrictions in place. On March 22 2020, the strictest level was put in place, where residents of Nova Scotia were asked to *Shelter-in-Place* and only essential services remained open. Our simulation model used these restrictions and the dates they were implemented to validate the model and test strategies [22].

**Table 2.** Restrictions levels that were implemented by the Nova Scotia government

| **Type** | **Level** | **Description** |
| --- | --- | --- |
| Closure | 1 | No closures |
|  | 2 | Mild restriction |
|  | 3 | Moderate restrictions |
|  | 4 | Severe restrictions |
|  | 5 | Shelter in place |
| Protection | 1 | No interventions |
|  | 2 | Masks required and physical distancing |
| Travel Restrictions | 1 | International travel |
|  | 2 | Domestic travel |
|  | 3 | Atlantic travel |
|  | 4 | No travel |

### Vaccination Campaign

A vaccination schedule was built into the model using Nova Scotia’s existing and planned vaccination rollout. The initial phases focused on frontline workers who had an increased likelihood of encountering infected individuals and residents of long-term care facilities. The vaccinations completed in the initial phases targeted people over 80 and healthcare workers aged 20-64. The last phase of the rollout is age-based only, starting with people over 80 and proceeding downward in 5-year increments. The entire eligible population is expected to have access to their first vaccine by June 30, 2021 [23]. We determined the minimum daily vaccinations by age group needed to meet the June deadline and modelled the resulting schedule. The second vaccine dose (or completed vaccine) will occur a maximum of 4 months later, which models the worst-case scenario. Our model has given a general vaccine efficacy of 85% at the first dose, and efficacy of 92% at the second dose which is an approximation of the vaccines that are currently being used in Nova Scotia (Pfizer-BioNTech, Moderna, AstraZeneca). We have also assumed that 90% of the population 12 years old and older will choose to get a vaccine. The model parameters used for the vaccination are provided in Table 3.

**Table 3.**
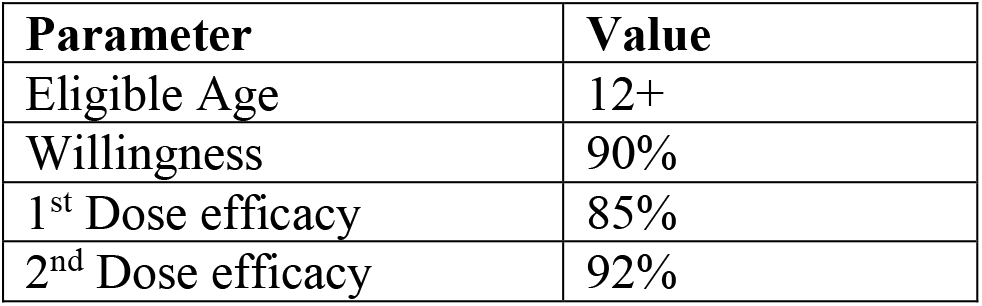
Model Parameters Used for Vaccination

### Model Parameters by Restriction Level

Both physical distancing as well as the use of non-medical masks have shown a reduction in transmissibility of the SARS-CoV-2 coronavirus [24]. Data from this study [24] were used for the transmissibility parameter in the model. For the importation numbers in our SEIR model, we used historical travel data. There are four different levels of travel policies in the model: No travel, Atlantic Canada, National, and International. The purpose of separating domestic travel into National and Atlantic was due to the implementation of the “Atlantic bubble”, an agreement among Atlantic Canadian provinces that permitted free movement between them without the need for isolation [25]. Statistics Canada provides data for the number of international and domestic travellers that visited Nova Scotia in 2017 and 2018 [26-31]. The domestic data was used to estimate the number of people that visit Nova Scotia from within Canada. A simplifying assumption was that the travellers are evenly distributed throughout the year. The yearly estimate was used to determine the number of incoming travellers per day. We also used this data to determine the number of trips that are essential for business for the various closure levels. The probability of infection of a traveller was determined based on the nation of origin. Table 4 shows the contact, transmissibility, and importation (due to travel) parameters that were used in the model.

**Table 4.** Model Parameters Used for Transmissibility, Contract Rate and Cases Imported.

| **Parameter Name** | **Level 1** | **Level 2** | **Level 3** | **Level 4** | **Level 5** | **Level 6** |
| --- | --- | --- | --- | --- | --- | --- |
| **Transmissibility** | 0.156 | 0.054 | 0.054 | 0.054 | 0.054 | 0.054 |
| **\*# Contacts / day** | 3.3 | 3.3 | 3.21 | 1.52 | 1.18 | 0.85 |
| **Case Imported / day (First Wave)** | 1.6359 | 1.4798 | 0.0965 | 0.0965 | 0 | 0 |
| **Case Imported / day (Second Wave)** | 16.359 | 14.798 | 0.965 | 0.965 | 0 | 0 |

\* Average contact rate shown. Model incorporated differences in contact rate by age.

### Modelling Software

The application of system dynamics simulation to SEIR models has been used extensively [13]. System dynamics are continuous simulation models that use stocks (compartments) and flows between them to represent processes of accumulation and feedback. The continuous-time dynamic model based differential equations is usually solved numerically by approximating it as a discrete-time model with small time increments. The model was developed using Vensim (ver 6.4, Ventana Systems Inc.; Harvard, MA, USA). The simulation was run and the results were exported into MS Excel (Microsoft LLC; Seattle, WA, USA) for graphing and comparison with actual values. The complete VENSIM model that we developed is shown on Figure 2.

**Figure 2:**
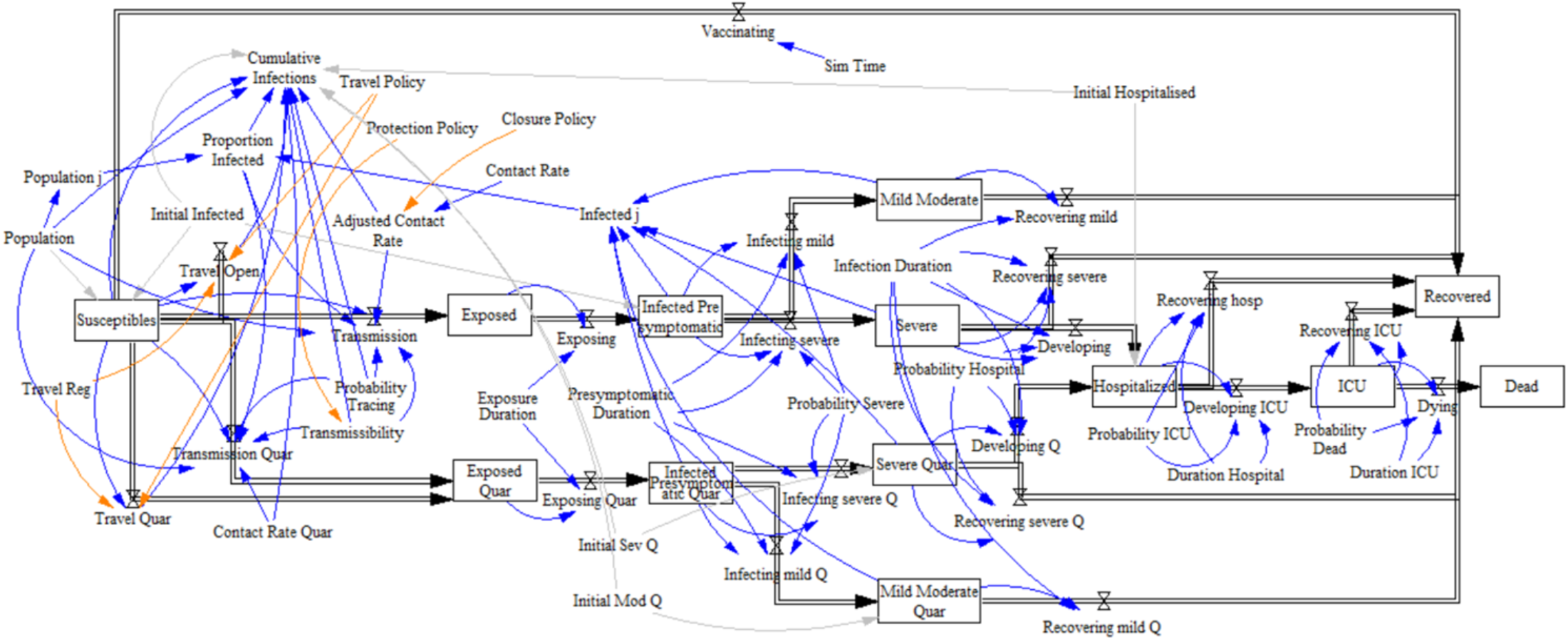
VENSIM Model of COVID-19 Infections in Nova Scotia

### Ethics

This study was approved by the research ethics board at NS Health. Study number 1025953.

## RESULTS

Initially, the simulation model was validated using publicly available data on the number of COVD-19 cases, recoveries and deaths in Nova Scotia, [32-34]. Figure 3 compares the model results to the active number of cases each day. Overall, the model closely follows the active cases. The actual data is skewed to the right of the model data, which is likely due the delay in symptom onset. This Figure is based on the start of the infections in Nova Scotia.

**Figure 3:**
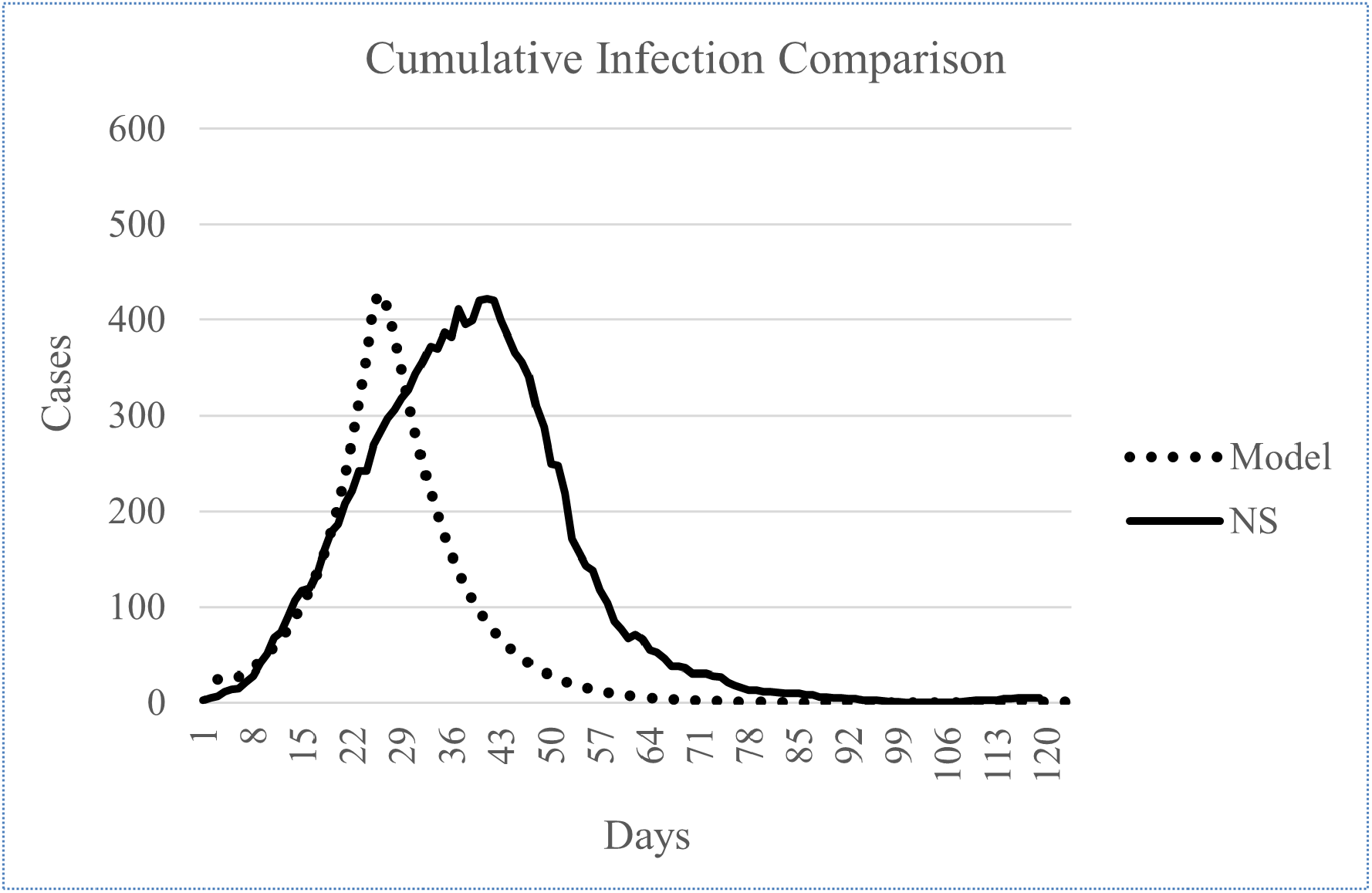
Model verification results for active cases in Nova Scotia compared to actual active cases reported. NS: Nova Scotia

### Results of Policy Scenarios

Several scenarios were run to determine the effects of changing policy restrictions on the COVID-19 infections, hospital admissions and deaths in Nova Scotia. These scenarios were run starting from March 15 2021 with each day shown after this date. Figure 4 shows the results of a conservative approach to the relaxation of restrictions. The top row, Figure 4.A, shows the results if there were no changes to the current restrictions in terms of closure level, continued use of masks and physical distancing, and a 2-week quarantine for travellers from outside of the province. This yields only a small number of active cases that is at the maximum at the start date. The middle row, Figure 4.B, shows the results with loosening of travel restrictions by allowing travel from Atlantic Canada without quarantine restrictions (Atlantic Bubble) on April 15 2021. This yields almost identical results as the top row. The bottom row goes further by loosening both closure and opening the Atlantic Bubble on April 15 2021, which yields only an additional 11 cases from option B. None of these scenarios resulted in any hospitalizations.

**Figure 4:**
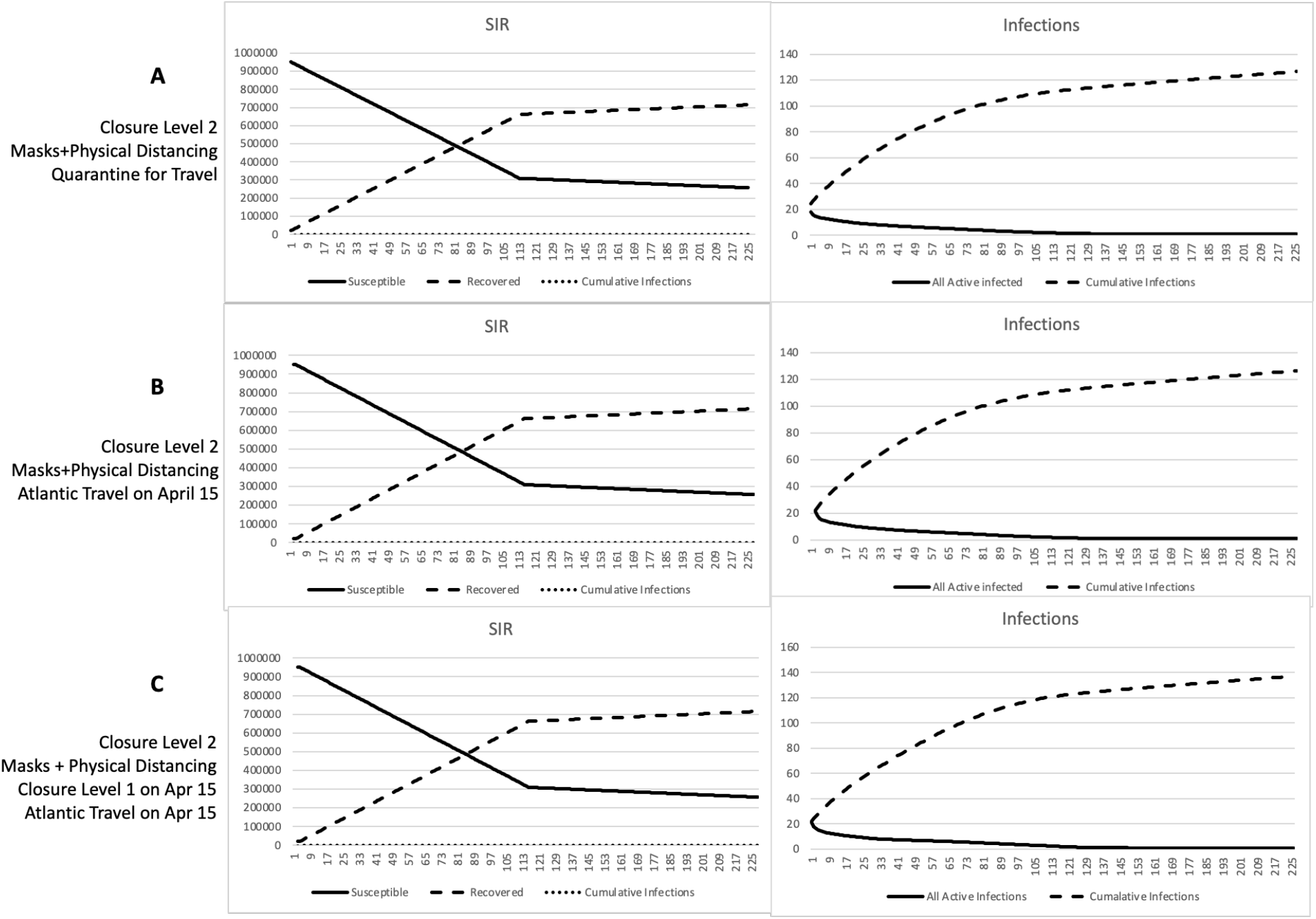
Model outputs with conservative policy scenarios from March 15 2021; the charts on the left show the change in the number of people susceptible and recovered; the charts on the right show the active and cumulative infections over time in Nova Scotia. A. Shows the change with no change in policy from March 15, 2021. B. Shows the change if the Atlantic Bubble opens on April 15 2021. C. Shows the change if the Atlantic Bubble opens and closure policy loosens on April 15. SIR=Susceptible Infected Recovered.

Figure 5 shows the effects of less restrictive policy options than the base case described above. The top row, Figure 5.A, shows the effects of opening the Atlantic Bubble on May 14, opening to national travel on July 1 2021, and moving to the least strict closure level on September 1 2021. This results in a steady increase in cases from July 9 2021 with 571 cumulative infections after one year; a small number of hospitalization results from this with less than 2 deaths over the year. The second row, Figure 5.B, shows the effects of opening the Atlantic Bubble on May 14, opening to national travel on July 5 2021, moving to the least strict closure level on September 3 2021, and toggling between removing and invoking the mask and physical distancing mandate from October 18 2021 onward. This results in a delayed infection of the remaining population that peaks on June 11 2022 with 5,975 active infections, and 102 hospitalizations on July 2 2022. The third row, Figure 5.C, shows the effects of opening the Atlantic Bubble and loosening closure policy on May 14, opening to national travel on July 5 2021, and removing the mask and physical distancing mandate from September 13 2021. This results in peak active infections of 6,114 by February 16, 2022 and 104 hospitalizations on March 8, 2022. The bottom row, Figure 5.D, shows the results from immediate opening of the Atlantic Bubble, the opening of all businesses without restrictions, and the removal of the mask and physical distancing mandate. This yields a maximum of 71,731active infections on June 4 2021 and a maximum of 1,097 hospitalizations on June 19 2021.

**Figure 5:**
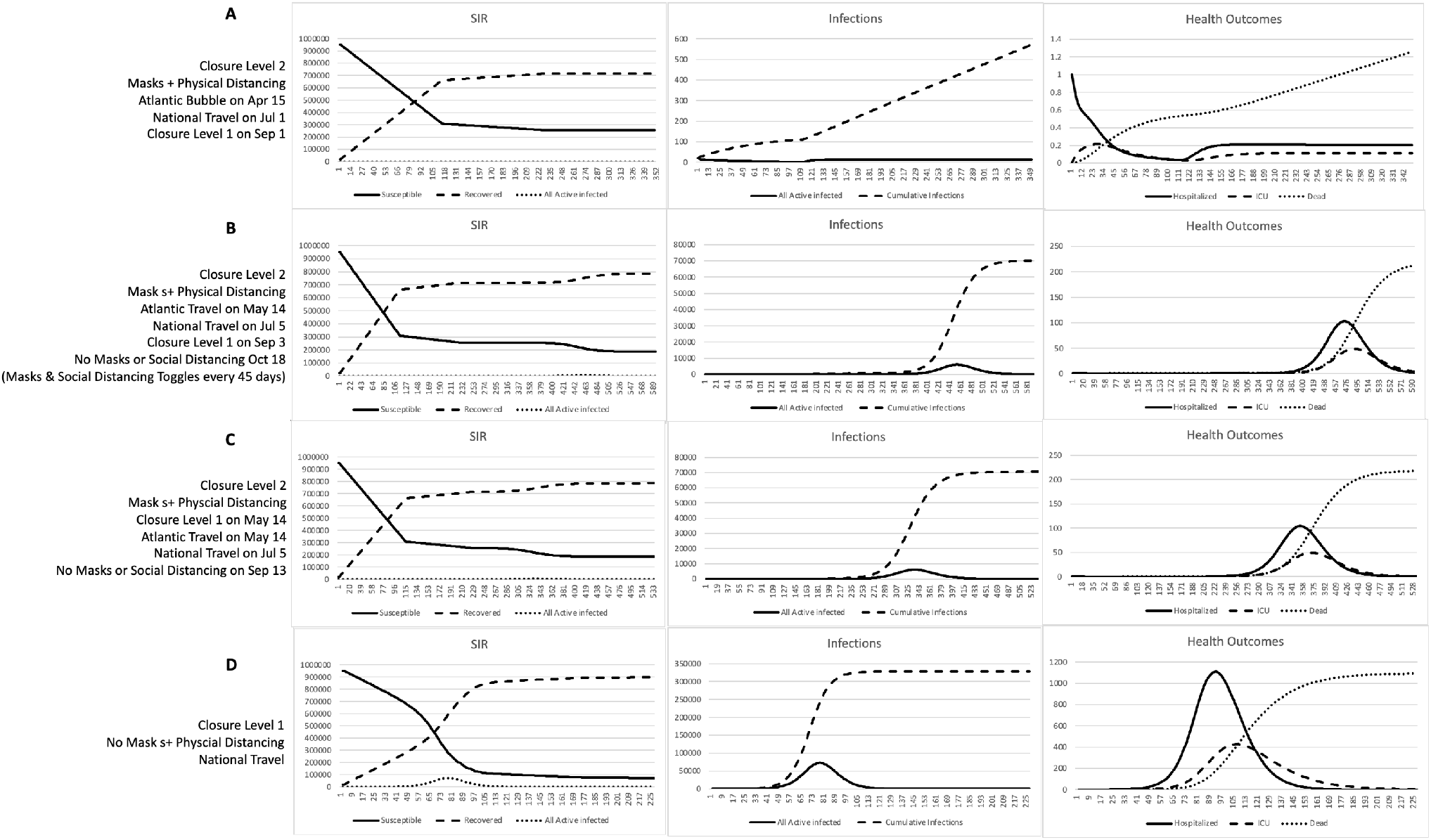
Model outputs with liberal policy scenarios from March 15 2021; the charts on the left show the change in the number of people susceptible and recovered; the charts on the middle show the active and cumulative infections; and the charts on the right show the hospitalizations, ICU admissions and deaths over time in Nova Scotia. A. Shows the change with the opening of the Atlantic Bubble on April 15 2021, allowing national travel on July 1 2021, and loosening the closure policy on September 1 2021. B. Shows the change with the opening of the Atlantic Bubble on May 15 2021, allowing national travel on July 5 2021, loosening of closure policy on September 3, 2021, and toggling every 45 days between removing and invoking the mask and physical distancing mandate beginning on October 18 2021. C. Shows loosening of closure policy and opening of Atlantic Bubble on May 14 2021, allowing national travel on July 1 2021, and removing the mask and physical distancing policy on September 6, 2021. D. shows the change if all closure, mask and physical distancing, and travel restrictions were removing immediately. SIR=Susceptible Infected Recovered; ICR=Intensive Care Unit.

## DISCUSSION

These results indicate that closure level, protection through masks and physical distancing, and restricting travel, all have significant effects on the number of COVID-19 infections and hospitalizations. By maintaining an Atlantic Bubble along with masks and physical distancing [34], Nova Scotia will continue to see a small number of infections even as restrictions on businesses and gatherings are loosened during the vaccination period. These results suggest that as long as the mask and physical distancing mandate is maintained while closure and travel restrictions are loosened, Nova Scotia will only see a small steady rise in the cumulative infections without any significant hospitalizations. However, if the mask and physical distancing mandate are removed by the fall, we will see the remaining unvaccinated population, drive a surge in infections. This will result in a peak of approximately 6,114 active infections, cumulative infections of over 70,000, and a peak of 104 hospitalizations by March of 2022. This is lower than the maximum impact if restrictions were to be removed immediately, which would result in over 70,000 active infections, 328,000 cumulative infections, and 1000 hospitalization.

These results show that although a herd immunity of 70% is required to quell infection rates [36], we will not reach this due to the combination of several factors. These factors include the assumed efficacy of the vaccines, the proportion of the population that will choose not to receive the vaccine, and not vaccinating children under the age of 12. For these reasons, these results indicate that masks and physical distancing will be required in order to continue to keep the case count and hospitalizations low, as closure and travel policies loosen. A slower opening, where the province is fully open by September and the maximum number of people are vaccinated results in a much smaller and manageable number of cases in early 2022. These results are possible due to Nova Scotia’s low vaccine hesitancy. Other provinces that are opening sooner may see a larger and more significant fourth wave.

## Supporting information

On-Line Supplemental

## Data Availability

Data used in this study is publicly available, and provided in citations in the manuscript

