## Supplementary material for "Effects of Various Policy Options on COVID-19 Cases in Nova Scotia including Vaccination Rollout Schedule: A Modelling Study": On-Line Supplemental

***Model Formulation***

The model formulation is provided fully in this section. The parameter definition is provided in the Table below. The change in the number of people that are susceptible over time is given by the following formula:

$$\frac{dS_{i,k}}{dt}=-\lambda_{i,k}S_{i,k}-\lambda_{q,i,k}S_{i,k}-\beta_{q}$$

The change in the number of people exposed over time is given by the following formula:

$$\frac{dE_{i,k}}{dt}=\left( 1-\sigma_{t} \right)\lambda_{i,k}S_{i,k}+\beta_{q}-\frac{1}{\sigma_{1}}E_{i,k}$$

The change in the pre-symptomatic infected population over time is given by the following formula:

$$\frac{d{IP}_{i,k}}{dt}=\frac{1}{\sigma_{1}}E_{i,k}-\frac{1}{\sigma_{2}}{IP}_{i,k}$$

The change in the infected population with mild to moderate symptoms is given by the following formula:

$$\frac{d{IM}_{i,k}}{dt}=\left( 1-\sigma_{sik} \right)\frac{1}{\sigma_{2}}{IP}_{i,k}-\frac{1}{\sigma_{3}}{IM}_{i,k}$$

The change in the infected population with severe symptoms is given by the following formula:

$$\frac{d{IS}_{i,k}}{dt}=\left( \sigma_{sik} \right)\frac{1}{\sigma_{2}}{IP}_{i,k}-\frac{1}{\sigma_{3}}{IS}_{i,k}$$

The change in the number of people who have been exposed over time but are in quarantine is given by the following formula:

$$\frac{d{EQ}_{i,k}}{dt}=\left( \sigma_{t} \right)\lambda_{q,i,k}S_{i,k}+\beta_{q}-\frac{1}{\sigma_{1}}{EQ}_{i,k}$$

The change in the infected pre-symptomatic population over time that is quarantined is given by the following formula:

$$\frac{d{IPQ}_{i,k}}{dt}=\frac{1}{\sigma_{1}}{EQ}_{i,k}-\frac{1}{\sigma_{2}}{IPQ}_{i,k}$$

The change in the infected population with mild to moderate symptoms over time that is quarantined is given by the following formula:

$$\frac{d{IMQ}_{i,k}}{dt}=\left( 1-\sigma_{sik} \right)\frac{1}{\sigma_{2}}{IPQ}_{i,k}-\frac{1}{\sigma_{3}}{IMQ}_{i,k}$$

The change in the infected population with severe symptoms over time that is quarantined is given by the following formula:

$$\frac{d{ISQ}_{i,k}}{dt}=\left( \sigma_{sik} \right)\frac{1}{\sigma_{2}}{IPQ}_{i,k}-\frac{1}{\sigma_{3}}{ISQ}_{i,k}$$

The change in the number of people hospitalized over time is given by the following formula:

$$\frac{dH_{i,k}}{dt}=\left( \sigma_{hik} \right)\frac{1}{\sigma_{3}}\left( {IS}_{i,k}+{ISQ}_{i,k} \right)-\frac{1}{\sigma_{4}}H_{i,k}$$

The change in the number of people in Intensive Care Units (ICU) is given by the following formula:

$$\frac{dI_{i,k}}{dt}=\left( \sigma_{c} \right)\frac{1}{\sigma_{4}}H_{i,k}-\frac{1}{\sigma_{5}}I_{i,k}$$

The number of people who die over time is given by the following formula:

$$\frac{dD_{i,k}}{dt}=\left( \sigma_{dik} \right)\frac{1}{\sigma_{5}}I_{i,k}$$

The number of people who recover over time given by the following formula:

$$\frac{dR_{i,k}}{dt}=\frac{1}{\sigma_{3}}\left( {IM}_{i,k}+{IMQ}_{i,k} \right)+\left( 1-\sigma_{hik} \right)\frac{1}{\sigma_{3}}\left( {IS}_{i,k}+{ISQ}_{i,k} \right)+\left( 1-\sigma_{c} \right)\frac{1}{\sigma_{4}}H_{i,k}+\left( 1-\sigma_{dik} \right)\frac{1}{\sigma_{5}}I_{i,k}$$

The force of infections that is used in the number of susceptible and exposed formula is given by the following formula:

$$\lambda_{i,k}= \gamma_{p}*c_{i,j,l}*\frac{\left( A_{i,k}+B_{i,k}+C_{i,k} \right)}{\sum_{i=1}^{I} \sum_{k=1}^{K} n_{i,k}}$$

The force of infections for those in quarantine is given by the following formula:

$$\lambda_{q,i,k}= \gamma_{p}*{cq}_{i,j,l} \frac{\left( F_{i,k}+G_{i,k}+H_{i,k} \right)}{\sum_{i=1}^{I} \sum_{k=1}^{K} n_{i,k}}$$

Table. Definition of Model Parameters

| **Parameter** | **Description** |
| --- | --- |
| $n_{i,k}$ | Population in age group i with comorbidity status k |
| $c_{i,j,l}$ | Contact rate |
| ${cq}_{i,j,l}$ | Quarantine contact rate |
| $\sigma_{1}$ | Average duration of exposed period |
| $\sigma_{2}$ | Average duration of pre-symptomatic period |
| $\sigma_{3}$ | Average duration of infection period |
| $\sigma_{4}$ | Average duration in hospital |
| $\sigma_{5}$ | Average duration in ICU |
| $\sigma_{t}$ | Probability exposed case is quarantined |
| $\sigma_{sik}$ | Probability case becomes severe |
| $\sigma_{hik}$ | Probability case is hospitalized |
| $\sigma_{c}$ | Probability case goes to ICU |
| $\sigma_{dik}$ | Probability of dying |
| $\beta_{q}$ | Cases imported under travel policy q |
| $\gamma_{p}$ | Transmissibility under protection policy p |
| $\lambda_{i,k}$ | Force of infection |
| ${\lambda_{q,i,k}}_{i}$ | Quarantine force of infection |
